# An analysis of SARS-CoV-2 viral load by patient age

**DOI:** 10.1101/2020.06.08.20125484

**Authors:** Terry C. Jones, Barbara Mühlemann, Talitha Veith, Guido Biele, Marta Zuchowski, Jörg Hofmann, Angela Stein, Anke Edelmann, Victor Max Corman, Christian Drosten

**Affiliations:** Institute of Virology, Charité-Universitätsmedizin Berlin, corporate member of Freie Universität Berlin, Humboldt-Universität zu Berlin, and Berlin Institute of Health, 10117 Berlin, Germany; Centre for Pathogen Evolution, Department of Zoology, University of Cambridge, Downing St., Cambridge, CB2 3EJ, U.K.; German Centre for Infection Research (DZIF), partner site Charité, 10117 Berlin, Germany; Norwegian Institute of Public Health, 0473 Oslo, Norway; Labor Berlin - Charité Vivantes GmbH, Sylter Straße 2, 13353 Berlin, Germany

## Abstract

As children are under-represented in current studies aiming to analyse transmission of SARS-coronavirus 2 (SARS-CoV-2), their contribution to transmission is unclear. Viral load, as measured by RT-PCR, can inform considerations regarding transmission, especially if existing knowledge of viral load in other respiratory diseases is taken into account. RT-PCR threshold cycle data from 3303 patients who tested positive for SARS-CoV-2 (out of 77,996 persons tested in total, drawn from across Germany) were analysed to examine the relationship between patient age and estimated viral load. Two PCR systems were used. In data from the PCR system predominantly used for community and cluster screening during the early phase of the epidemic (Roche LightCycler 480 II), when such screening was frequent practice, viral loads do not differ significantly in three comparisons between young and old age groups (differences in log_10_ viral loads between young and old estimated from raw viral load data and a Bayesian mixture model of gamma distributions collectively range between −0.11 and −0.43). Data from a second type of PCR system (Roche cobas 6800/8800), introduced into diagnostic testing on March 16, 2020 and used during the time when household and other contact testing was reduced, show a credible but small difference in the three comparisons between young and old age groups (differences, measured as above, collectively range between −0.43 and −0.83). This small difference may be due to differential patterns of PCR instrument utilization rather than to an actual difference in viral load. Considering household transmission data on influenza, which has a similar viral load kinetic to SARS-CoV-2, the viral load differences between age groups observed in this study are likely to be of limited relevance. Combined data from both PCR instruments show that viral loads of at least 250,000 copies, a threshold we previously established for the isolation of infectious virus in cell culture at more than 5% probability, were present across the study period in 29.0% of kindergarten-aged patients 0-6 years old (n=38), 37.3% of those aged 0-19 (n=150), and in 51.4% of those aged 20 and above (n=3153). The differences in these fractions may also be due to differences in test utilization. We conclude that a considerable percentage of infected people in all age groups, including those who are pre- or mild-symptomatic, carry viral loads likely to represent infectivity. Based on these results and uncertainty about the remaining incidence, we recommend caution and careful monitoring during gradual lifting of non-pharmaceutical interventions. In particular, there is little evidence from the present study to support suggestions that children may not be as infectious as adults.

## Introduction

Measures to curb the spread of SARS-CoV-2 by non-pharmaceutical interventions are beginning to show effects in many countries. Along with the gradual lifting of measures of physical distancing, there is a growing discussion regarding the contribution of school and kindergarten closures to the reduction of transmission rate (*1*) and to the expected rebound from reopening. Studies to determine the contribution of children as sources of infection are complicated by the fact that non-pharmaceutical interventions, including school and kindergarten closures, were in place before observational trials could begin. Studies on primary and secondary attack rates suggest that children could be infected by SARS-CoV-2 at a rate that may not be different from that of adults (*2–6*). However, considerable uncertainty remains regarding the influence of different contact behaviour in children versus adults, and the extent to which children can act as sources of infection in general. A challenge when trying to address this question by epidemiological observation is posed by the present situation of physical distancing. Because kindergartens and schools have been closed, it becomes less likely that children become index cases in households. During the early phase of the SARS-CoV-2 epidemic in many European countries, the seeding of cases by adult-aged travelers who visited early epidemic foci was an additional reason why children were under-represented in age-related incidence (*7*). It is therefore unlikely that epidemiological investigations undertaken under the present conditions can identify the actual risk of acquisition of infection from children by subjects of any age group.

An alternative way to obtain a proxy of infectivity is to analyze virus concentration in the respiratory tract (henceforth referred to as ‘viral load’) and compare this with knowledge of transmission in related diseases for which data regarding the relationship between viral load and infectivity are available. We have previously shown that viral loads, expressed as RNA copies per mL of sample or entire swab specimen, predict the likelihood of isolation of infectious virus in cell culture (*8*). Probit regression analysis predicted that samples with a log_10_ viral load below 5.4 (∼250,000 copies per mL) have a probability of yielding a virus isolate below 5%. We also found that virus could not be isolated from respiratory samples after the first week of symptoms, which is concordant with transmission analyses based on actual transmission pairs, suggesting that most transmission stops by the end of the first week of symptoms (*8*). To enable an estimate of infectivity in children, we analyzed viral loads observed during routine testing at a large diagnostic laboratory in Berlin (Charité Institute of Virology and Labor Berlin). Charité Institute of Virology was the first laboratory qualified to test for SARS-CoV-2 in Germany, and until early February 2020 was the only SARS-CoV-2 testing facility in Berlin, a city of ca. 3.8 million inhabitants. Labor Berlin is a large medical laboratory services provider in Berlin, owned by the senate of Berlin and serving Charité as well as other large hospitals in Berlin and beyond. Labor Berlin serves public testing centres that mainly see adult outpatients. It also tests out- and in-patients from several hospitals, and serves practitioners and public health agencies submitting samples taken during contact tracing.

## Results

From January to May 2020, virology laboratories at Charité and Labor Berlin screened 77,996 patients for SARS-CoV-2 infection. Of these, 3,303 (4.2%) had at least one positive result. Initial testing exclusively used Roche LightCycler 480 II (LC480) PCR instruments. From mid-March, Roche cobas 6800 and 8800 (cobas) instruments were introduced to increase testing capacity (**Figure 1**). Positive PCR results were obtained in 1382 of 26,453 (5.2%) tests on LC480 and 2216 of 67,548 (3.3%) tests on cobas instruments. The per-instrument totals sum to more than the patient total due to some patients being tested and testing positive multiple times on both systems. The difference in instrument detection rates is due to differences in test and instrument utilization, as explained below.

**Figure 1:**
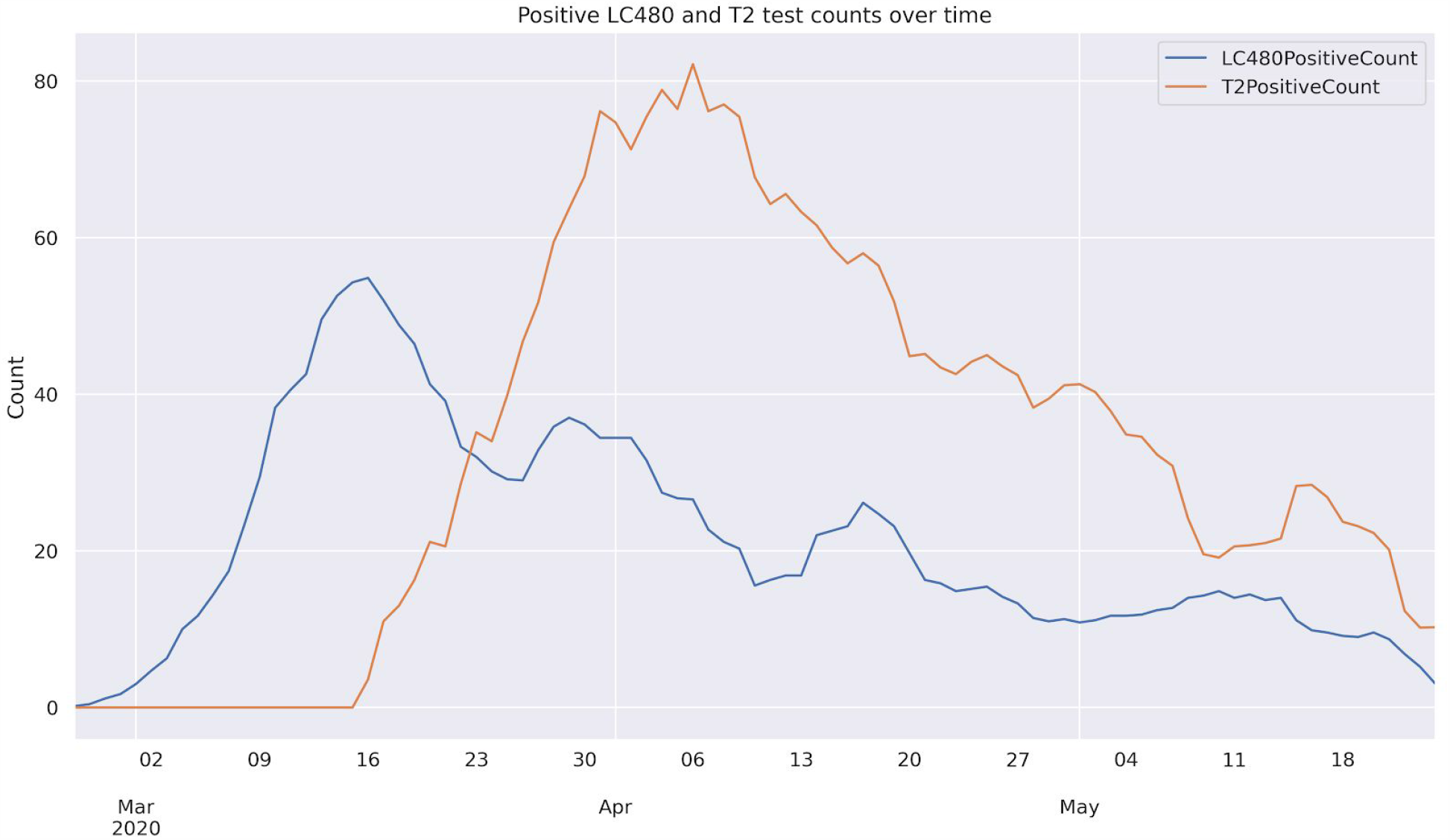
Utilization of LC480 and cobas test systems over the study period. ‘LC480Count’, in blue, shows the number of tests performed using the Roche LightCycler II (LC480) machine, ‘T2Count’, in orange, shows tests performed on the cobas machines over time. Six day moving average.

### Test utilization

To obtain an impression of test utilization, we identified, among all submitting clinical entities, a group of outpatient departments and practices that typically see early mild cases for initial infection screening. This includes a community testing centre established by Charité for the general public in a central location in Berlin, the Charité department of travel medicine outpatient centre, and public health agencies in Berlin city districts and regions outside Berlin submitting samples from transmission cluster investigations. Results from this group of submitting entities are hereafter referred to as “community/cluster testing” results. We observe a striking difference of the fraction of community/cluster testing in March compared to April and May. Community/cluster testing decreases from ca. 14% to ca. 7% for children, and ca. 21% to ca. 6% in adults (**Table 1**). This shift coincides with the incidence peak of SARS-CoV-2 infections.

**Table 1:** Test utilization in March, April, and May 2020.

| **Children (0-9 years)** | **March** | **April** | **May** |
| --- | --- | --- | --- |
| All | 1,521 | 1,125 | 1,360 |
| Community/cluster testing* | 210 (13.8%) | 82 (7.2%) | 96 (7.1%) |
| **Adults (>20 years)** |  |  |  |
| All | 24,414 | 46,389 | 39,868 |
| Community/cluster testing* | 5255 (21.5%) | 3080 (6.6%) | 2062 (5.2%) |

\*Samples from outpatient departments that typically see early, mild cases for initial infection screening. This also includes community testing centres as well as public health authorities submitting samples from cluster investigations. The figures for May 2020 are through the 24th.

This pattern can be explained by two effects. First, the lower fraction of children tested in community/cluster testing may be due to the fact that children are less likely to be tested due to milder (or no) symptoms as compared to adults. They are also less likely to be presented at overcrowded community testing centres by their parents. Second, in later stages of the outbreak, public health agencies submitted fewer samples from cluster contact tracing, which often includes mildly- or asymptomatic children.

In addition, multiple factors led to the preferential testing of certain patient groups on either the LC480 or the cobas testing system. The LC480 system was the only one available before contact cluster testing declined toward the end of March (**Figure 1**). The viral loads measured using this test system thus better reflect patients with early symptoms or pre-symptomatic patients typically seen in transmission cluster studies or medical practices. An important aspect of test utilization is the fact that the cobas system uses a special sample buffer for sample collection. This led to preferential cobas system testing of samples originating from clinical centres closely collaborating with Labor Berlin with this sample buffer available, influencing the age mix of the patient population tested on that instrument. First, cobas samples from community testing are primarily from adult patients, as community samples tested on the cobas system mainly included the Charité community testing centre and the Charité travel medicine outpatient department, both of which have more adults than pediatric patients. In contrast, the generic throat swab specimens collected by practitioners or health agencies performing household contact testing are mainly processed on the LC480 system that does not require the specialized sample buffer. Additionally, hospitalized pediatric patients (as opposed to children from household testing cases) are biased towards the cobas testing system, as the treating departments are provided with the cobas sampling buffer. As hospitalization occurs later in the course of infection, the viral loads in these patients are lower than in outpatients. This effect is confirmed for the young age groups in our data (**Figure 2**). The statistically significant higher proportion of high log_10_ viral load results from LC480 in community/cluster testing for children is shown in **Figure 3** (Fisher’s two-tailed exact test, p-values: 0.015 and 0.039 for those aged 0-9 and 0-19, respectively).

**Figure 2:**
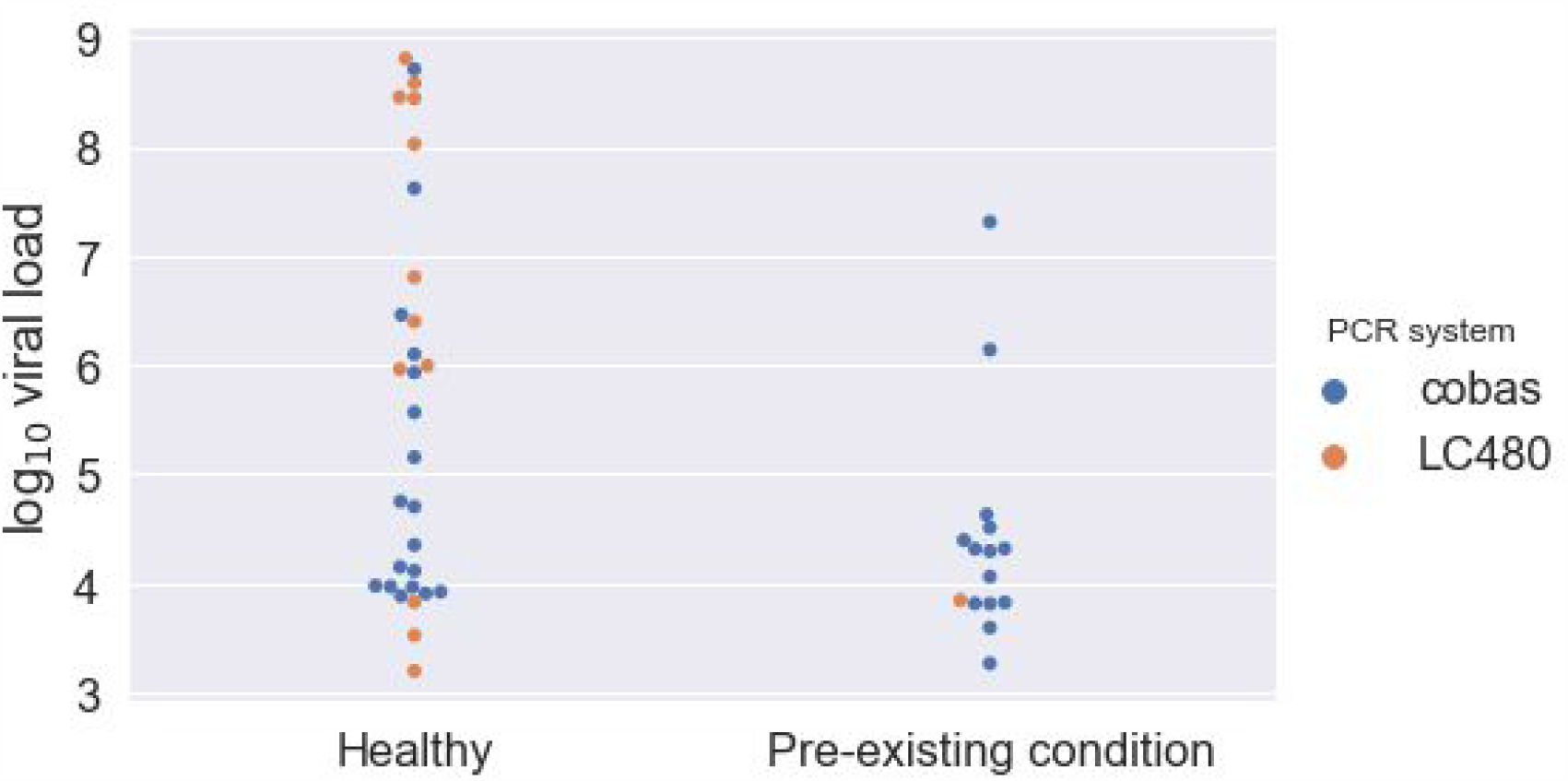
Differences in viral load in an exemplary group of patients aged 0-11 years with and without a pre-existing condition. Mean log_10_ viral loads in the healthy and the pre-existing condition categories are 5.642 and 4.408, respectively. Parametric (Welch’s t-test) and non-parametric (Mann-Whitney rank test) find a significant difference in viral loads between children that are healthy and those with a pre-existing condition (p-values 0.027 and 0.006, respectively), when testing the combined viral loads from both PCR systems. When only considering viral loads measured by cobas instruments, no significant difference is detected.

**Figure 3:**
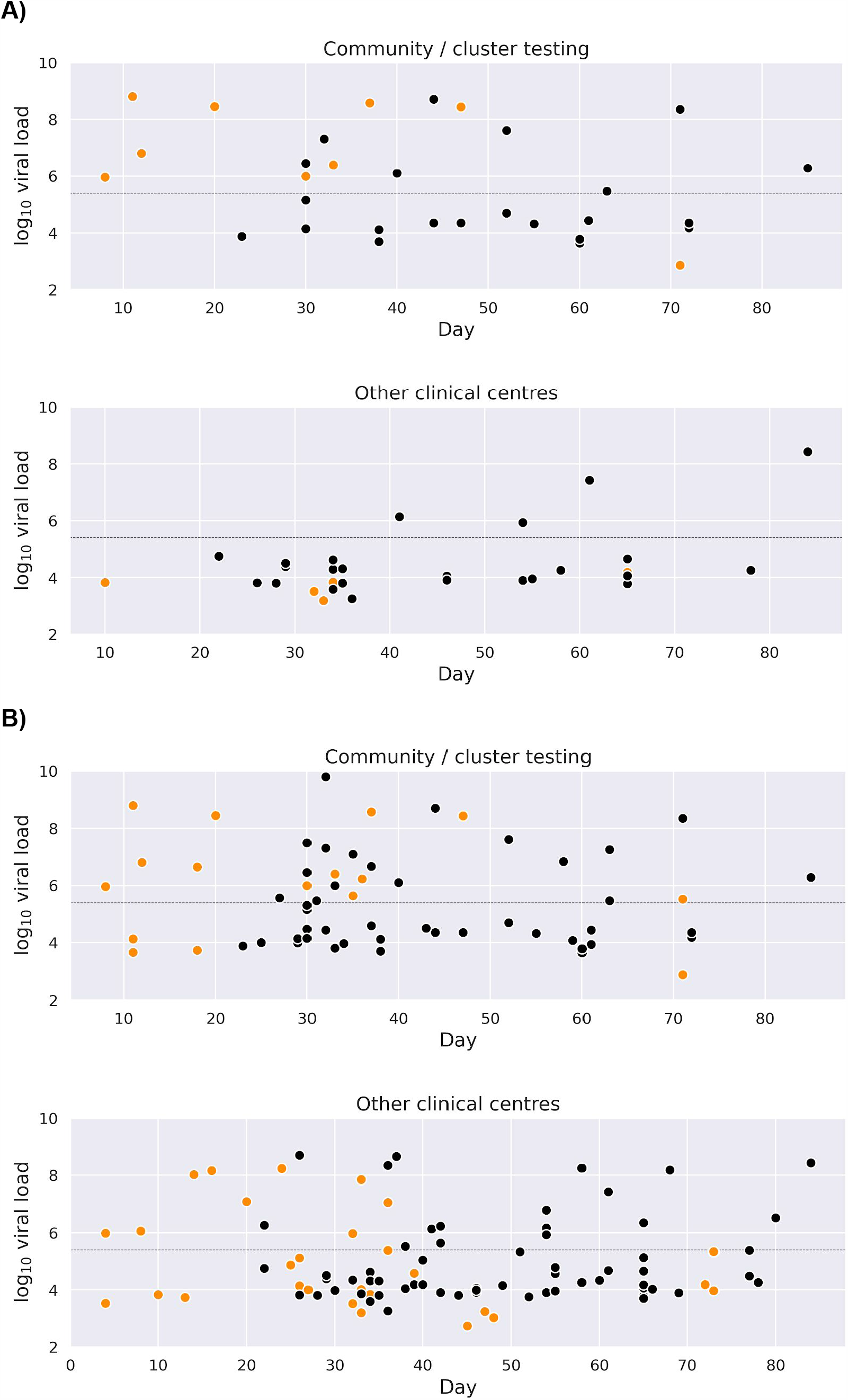
Viral loads over time in children from **A)** 0-9 and **B)** 0-19 year olds. Orange data points indicate tests done on LC480 machines, black on cobas. Community/cluster testing includes children tested at walk-in clinics, as well as those tested due to having contact with a SARS-CoV-2 positive individual, asymptomatic cases, and family clusters. ‘Other clinical centres’ includes children tested in hospitals. The dotted line denotes a log_10_ viral load of 5.4, corresponding to 250,000 viral copies. Means and standard deviations of log_10_ viral loads in household/cluster and other testing centres can be found in **Table 5**. In those aged 0-9 (**Figure A**), 31 children were tested in household/cluster testing (9 on LC480, 22 on cobas instruments), 30 children were tested in other clinical centres (6 on LC480, 24 on cobas instruments). Two children tested on both LC480 and cobas instruments are included in both counts; both those children were tested in community/cluster testing. In community/cluster testing, 52% of children had viral loads higher than 250,000 viral copies, in other clinical centres only 13%. In community/cluster testing, significantly more of the children with log_10_ viral loads above 250,000 were tested on LC480 but the same is not the case for other testing centres (Fisher’s two-tailed exact test p-values, 0.015 and 1.0 respectively). For those aged 0-19 (**Figure B**), 60 children were tested in household/cluster testing (16 (27%) on LC480 and 44 (73%) on cobas instruments), 94 in other clinical centres (32 (34%) on LC480 and 62 (66%) on cobas instruments). Four children were tested on LC480 and cobas instruments, two in community/cluster testing, and two in other clinical centres. In community/cluster testing, 50% of children had viral loads higher than 250,000 viral copies, in other clinical centres only 28%. As with age range 0-9 years (just mentioned), in community/cluster testing, significantly more of the children with log_10_ viral loads above 250,000 were tested on LC480 but the same is not the case for other testing centres (Fisher’s two-tailed exact test p-values, 0.039 and 1.0 respectively).

Finally, we observed a clear difference in the general structure of the threshold cycle (Ct) data reported by the LC480 and the cobas instruments (**Figure 4**). The LC480 provides Ct values that are evenly distributed across the entire viral load range of ca. 10^3^ to 10^11^ copies per swab or mL. For unknown technical reasons, the distribution of data from the cobas system exhibits an increased density between cycles 34 to 39, corresponding to viral loads between 5.0 and 3.5 (**Figure 4**). This difference suggests the data from the two instrument types should be evaluated and assessed separately in numerical comparisons that involve lower viral loads.

**Figure 4:**
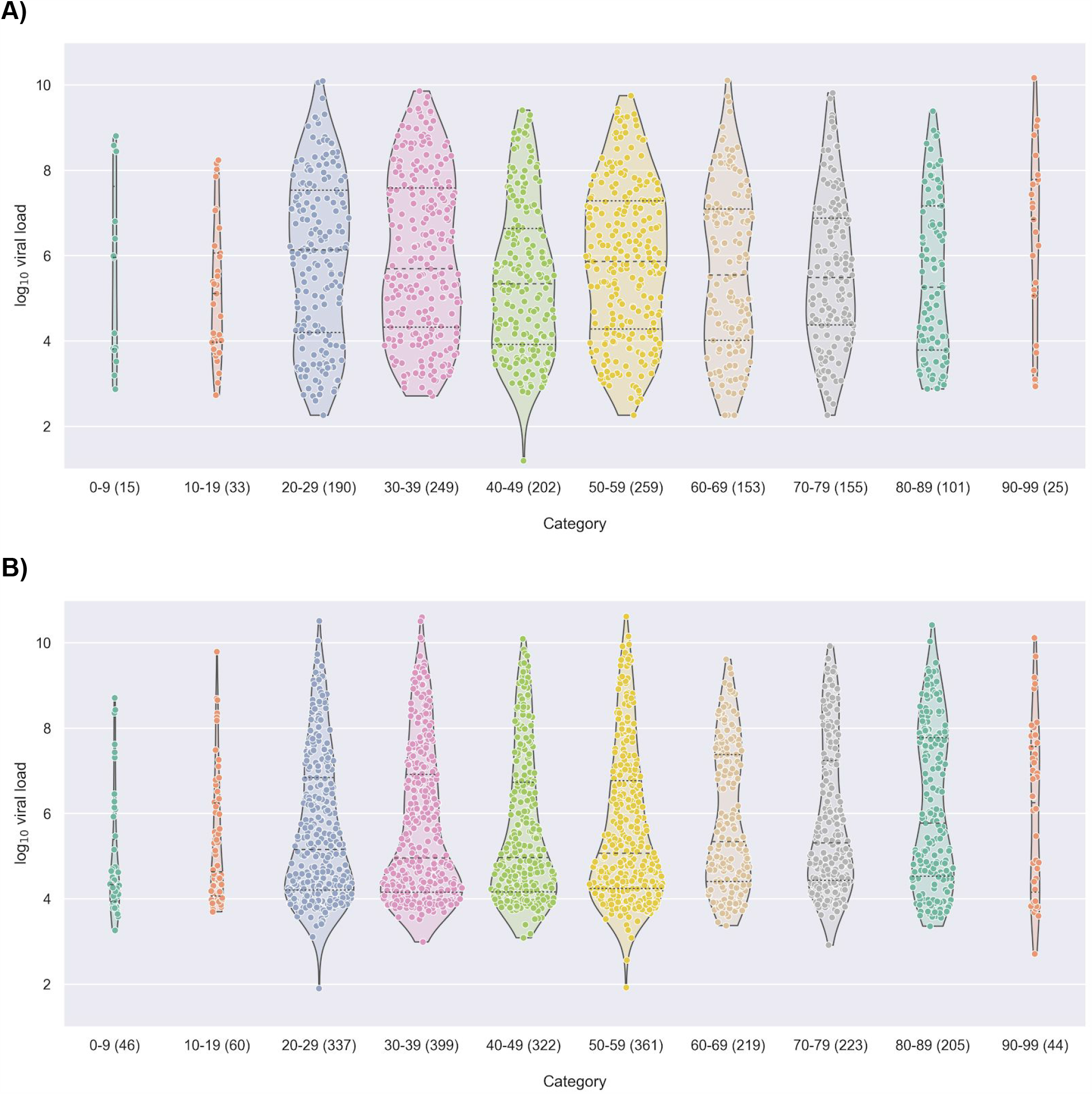
Distribution of viral loads by age group and PCR instrument. **A)** LC480, **B)** cobas. Note the pronounced additional density in the cobas system at log level ∼3.5-5.0 and the relative paucity of data at levels approaching 2.0 as compared to the LC480, as discussed in the main text and as shown in **Figure 6**.

### Statistical approach

To look for relationships between viral load and age, we took two approaches: 1) testing for statistical differences between the viral loads in aggregated age categories, and 2) treating age as a continuous variable and using a gamma regression to predict viral load. In the age categorization we made comparisons between three groups of patients: a) 0-9 years versus 10-99 years, b) 0-9 years versus 19-99 years, and c) 0-19 years versus 20-99 years. Age category viral loads were compared via the Mann-Whitney rank test and Welch’s t-test. The categories were also examined via a Bayesian analysis using gamma mixture models to account for the multi-modal nature of the viral load data. The Bayesian analysis used a mixture of three gamma distributions and accounted for variations between age groups by estimating age-group specific component weights. To provide a fine-grained overview of the data, log_10_ viral loads for an overall 10-year age bracket breakdown, with number and percentage of RT-PCR positive patients are shown in **Table 2**, while **Figures 4** and **5** show the distribution of viral load values in these groups.

**Table 2:**
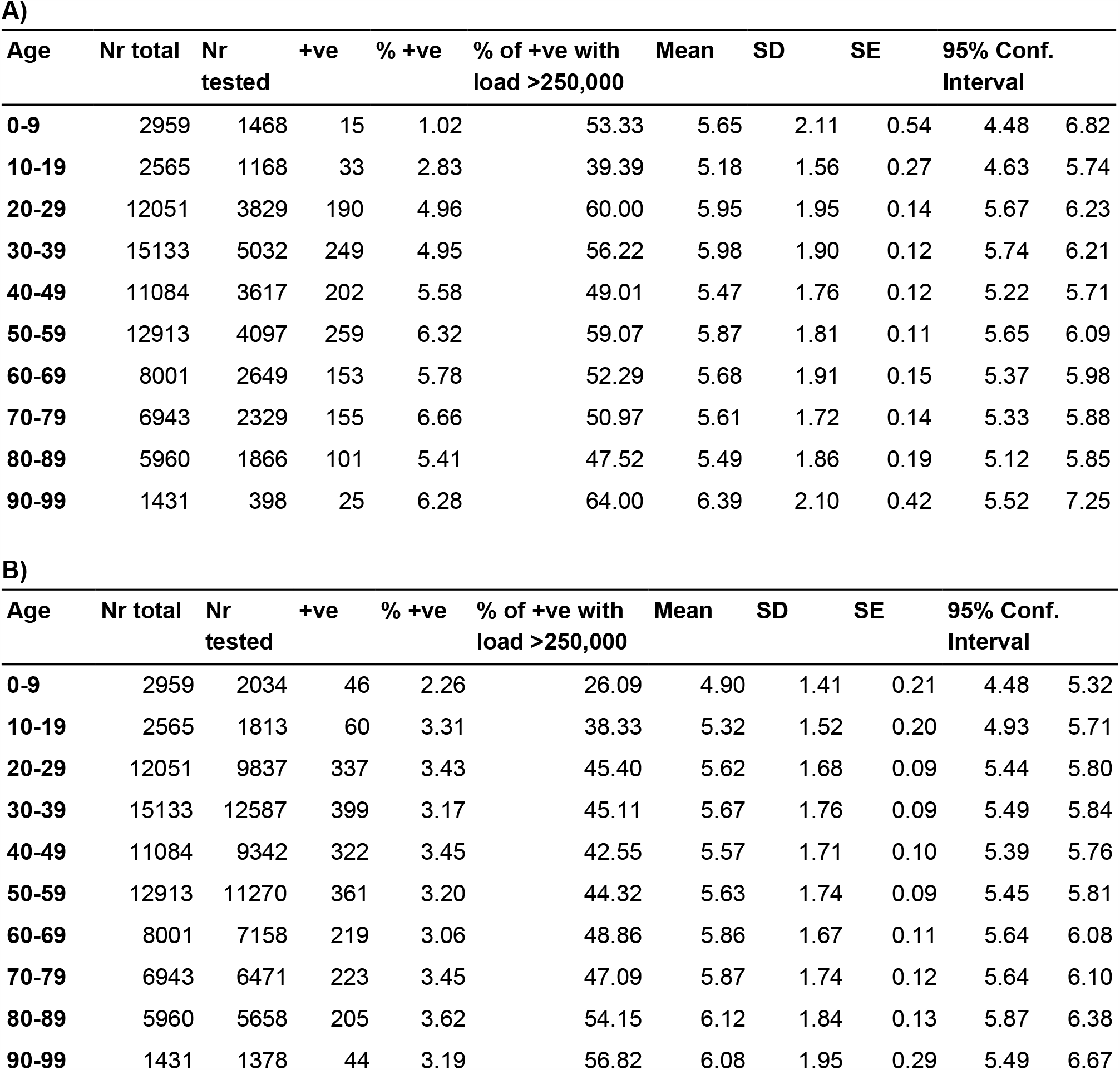
10-year age stratification showing positive PCR counts and percentages, and statistics describing viral load distributions. **A)** LC480 **B)** cobas. The ‘Nb total’ column in each categorization gives the total number of patients, the ‘Nb tested’ the number of patients tested with A) or B). ‘+ve’ indicates the total number of positive RT-PCR results for the subgroup. ‘% +ve’ is the percentage of tested people with a positive test result. ‘% of +ve with load >250,000’ indicates the percentage of the positively tested individuals with a viral load of over one million viral copies. Mean, standard deviation (SD), standard error (SE), 95% Confidence Interval (95% Conf.), and the interval are shown for the base 10 logarithm of viral load. +ve: positive, Nr: number.

**Figure 5:**
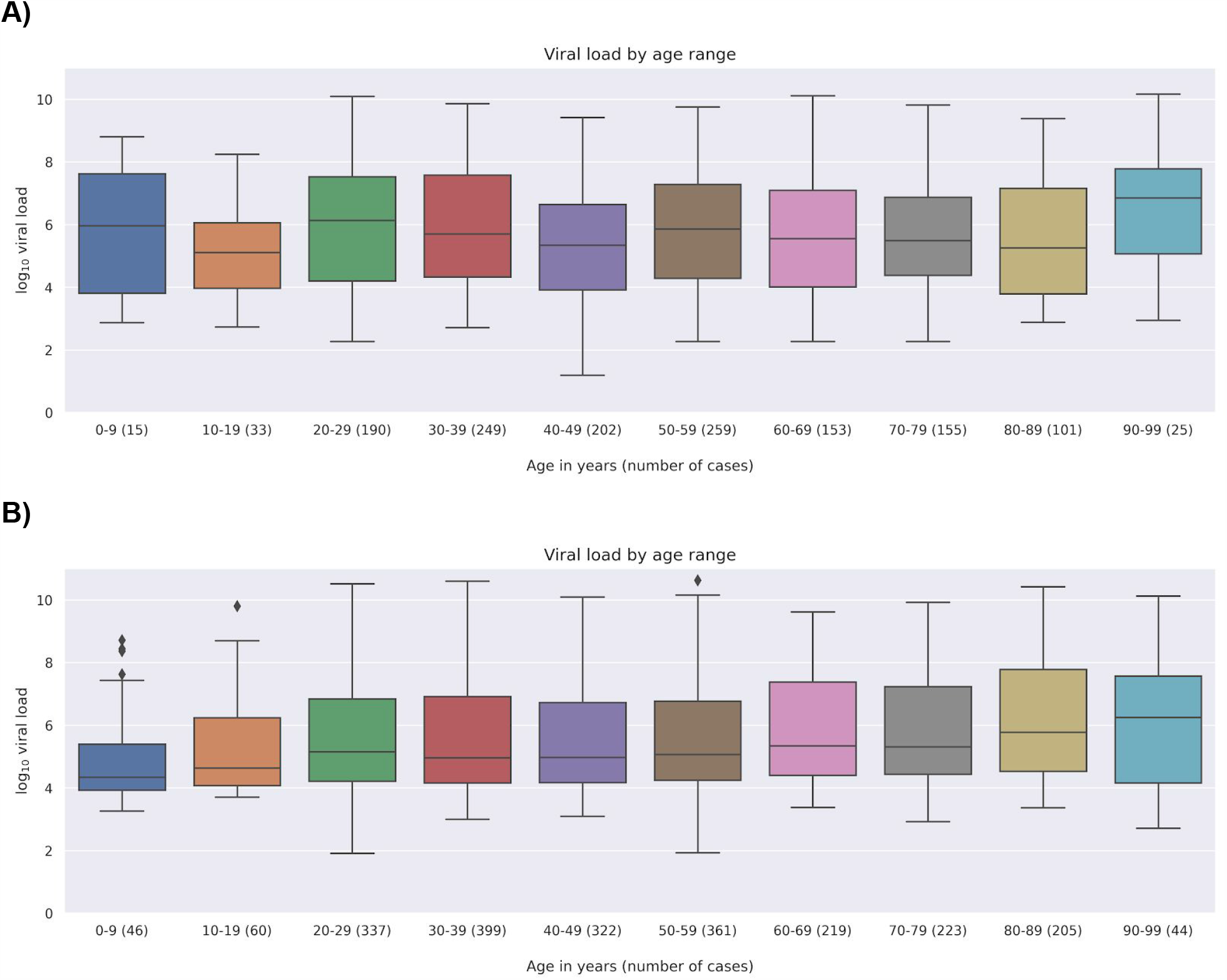
Viral load by patient 10-year age strata. The base 10 logarithm of viral load is estimated from the real-time PCR Ct value (see Methods). Counts of people in each age category are given in parentheses in the x-axis labels. **A)** LC480, **B)** cobas.

### Pairwise analysis of variation in viral load between age categories

We performed a parametric (using the Welch’s T-test) and non-parametric (using the Mann-Whitney rank test) comparison between age categories. We find no significant difference between any comparison involving data from the LC480 system, but do find small but significant differences between all three pairs of groups in the data from the cobas system (**Table 3**).

**Table 3:** A) Welch’s t-test, B) Mann-Whitney rank test for the difference in means and medians (respectively) between the three age group splits.

| PCR | Comparison | N | Means | Difference in means | SE of difference in means | 95% CI for difference in means | p-value | Statistic |
| --- | --- | --- | --- | --- | --- | --- | --- | --- |
| LC480 | 0-9 vs >9 | 15 / 1367 | 5.646 / 5.754 | -0.108 | 0.566 | -1.321 to 1.105 | 0.852 | -0.190 |
|  | 0-9 vs >19 | 15 / 1334 | 5.646 / 5.768 | -0.122 | 0.566 | -1.335 to 1.091 | 0.833 | -0.215 |
|  | 0-19 vs >19 | 48 / 1334 | 5.328 / 5.768 | -0.440 | 0.262 | -0.965 to 0.086 | 0.099 | -1.679 |
| cobas | 0-9 vs >9 | 46 / 2170 | 4.902 / 5.722 | -0.820 | 0.231 | -1.249 to -0.391 | 0.000 | -3.843 |
|  | 0-9 vs >19 | 46 / 2110 | 4.902 / 5.733 | -0.831 | 0.213 | -1.261 to -0.402 | 0.000 | -3.895 |
|  | 0-19 vs >19 | 106 / 2110 | 5.138 / 5.733 | -0.595 | 0.150 | -0.892 to -0.298 | 0.000 | -3.965 |

**B)**
| PCR | Comparison | N | Medians | Median of pairwise differences | 95% CI for median of pairwise differences | p-value | Statistic |
| --- | --- | --- | --- | --- | --- | --- | --- |
| LC480 | 0-9 vs >9 | 15 / 1367 | 5.961 / 5.664 | -0.155 | -0.196 to -0.101 | 0.783 | 9828 |
|  | 0-9 vs >19 | 15 / 1334 | 5.961 / 5.688 | -0.169 | -0.211 to -0.113 | 0.764 | 9555 |
|  | 0-19 vs >19 | 48 / 1334 | 5.218 / 5.688 | -0.434 | -0.458 to -0.407 | 0.115 | 27736 |
| cobas | 0-9 vs >9 | 46 / 2170 | 4.334 / 5.150 | -0.595 | -0.607 to -0.580 | 0.000 | 34634 |
|  | 0-9 vs >19 | 46 / 2110 | 4.334 / 5.156 | -0.604 | -0.618 to -0.592 | 0.000 | 33507 |
|  | 0-19 vs >19 | 106 / 2110 | 4.460 / 5.156 | -0.429 | -0.438 to -0.417 | 0.000 | 88415 |

Differences in mean log_10_ viral loads (mean in the young minus mean in the older) for the Welch’s t-test are as follows: 0-9 vs >9 years: −0.108 and −0.820; 0-9 vs >19 years: −0.122 and −0.831; 0-19 vs >19: −0.44 and −0.595 for the LC480 and cobas systems, respectively. For the Mann-Whitney rank test, the corresponding difference in medians are: 0-9 vs >9 years: −0.155 and −0.595; 0-9 vs >19 years: −0.169 and −0.604; 0-19 vs >19: −0.434 and −0.429 for the LC480 and cobas systems, respectively.

A Bayesian analysis modelling log_10_ viral loads as a mixture of gamma distributions in Stan (*9, 10*) found a difference of −0.61 (−1.12, 0.00) log_10_ viral loads for the comparison of the youngest age group (0-9) against those aged >9 years, −0.62 (−1.14, −0.01) for the comparison between 0-9 and >19, and −0.47 (−0.87, −0.02) for the comparison of those aged 0-19 against those aged >19 years in the cobas data. In the LC480 data, the credible interval for the difference between means contained zero in all three cases, with the estimated differences being −0.2 (−1.51, 1.2), −0.21 (−1.53, 1.19), and −0.42 (−1.1, 0.31), respectively **(Table 4)**. Numbers in parentheses indicate the 95% credible intervals.

**Table 4:** Estimated differences in log_10_ viral loads between groups using a mixture model of gamma distributions.

| PCR | Comparison | $\log_{10}$ viral load difference (95% credible interval; posterior probability, given the model and data, that the difference is less than zero) |
| --- | --- | --- |
| LC480 | 0-9 vs >9 | -0.2 (-1.51, 1.2; 0.61) |
|  | 0-9 vs >19 | -0.21 (-1.53, 1.19; 0.62) |
|  | 0-19 vs >19 | -0.42 (-1.1, 0.31; 0.88) |
| cobas | 0-9 vs >9 | -0.61 (-1.12, 0; 0.98) |
|  | 0-9 vs >19 | -0.62 (-1.14, -0.01; 0.98) |
|  | 0-19 vs >19 | -0.47 (-0.87, -0.02; 0.98) |

**Table 5:**
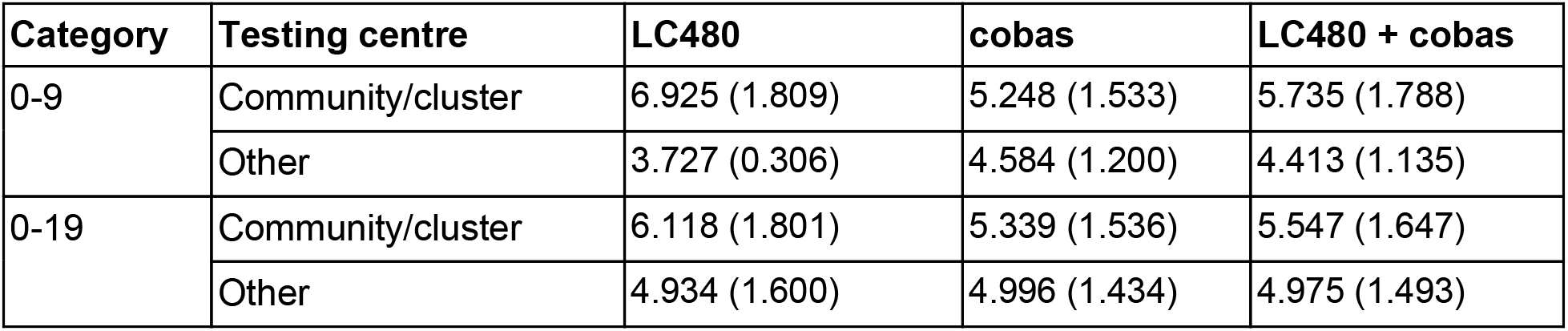
Means and standard deviations of log_10_ viral loads in household / community and other testing centres.

### Analysis using age as a continuous variable

We performed a Bayesian gamma regression using the brms package (*11, 12*) in R (*13*), predicting viral load from age, type of PCR system (LC480 or cobas), and an age-PCR system interaction. In the following text, numbers in parentheses indicate the 95% credible intervals. We found a small positive association between age and viral load (0.18 (0.11, 0.25)) log_10_ viral load increase per 1 SD increase in age for the cobas data, and a small negative association (−0.06 (−0.17, 0.04)) for the LC480 data (**Figure 6**). The same analysis found, for the cobas sample, a difference of −0.38 (−0.84, 0.1) log_10_ viral loads for the comparison of the youngest age group (0-9 years) with those aged >9 years, −0.39 (−0.85, 0.09) for the comparison of the youngest group with those over 19, and −0.34 (−0.65, −0.02) for the comparison of the 0-19 year group with those aged >19 years. In the LC480 data, the results for these comparisons are, in the same order, 0.14 (−0.87, 1.26), 0.15 (−.85, 1.22), and 0.13 (−0.44, 0.73) log_10_ viral loads in those aged 0-19 vs >19 years old. A gamma regression using splines (not shown) to allow a non-linear relationship between age and log_10_ viral load found consistent results, with a minor difference being that the age-viral load association was also positive for the LC480 sample, wherein credible intervals overlapped with zero.

**Figure 6:**
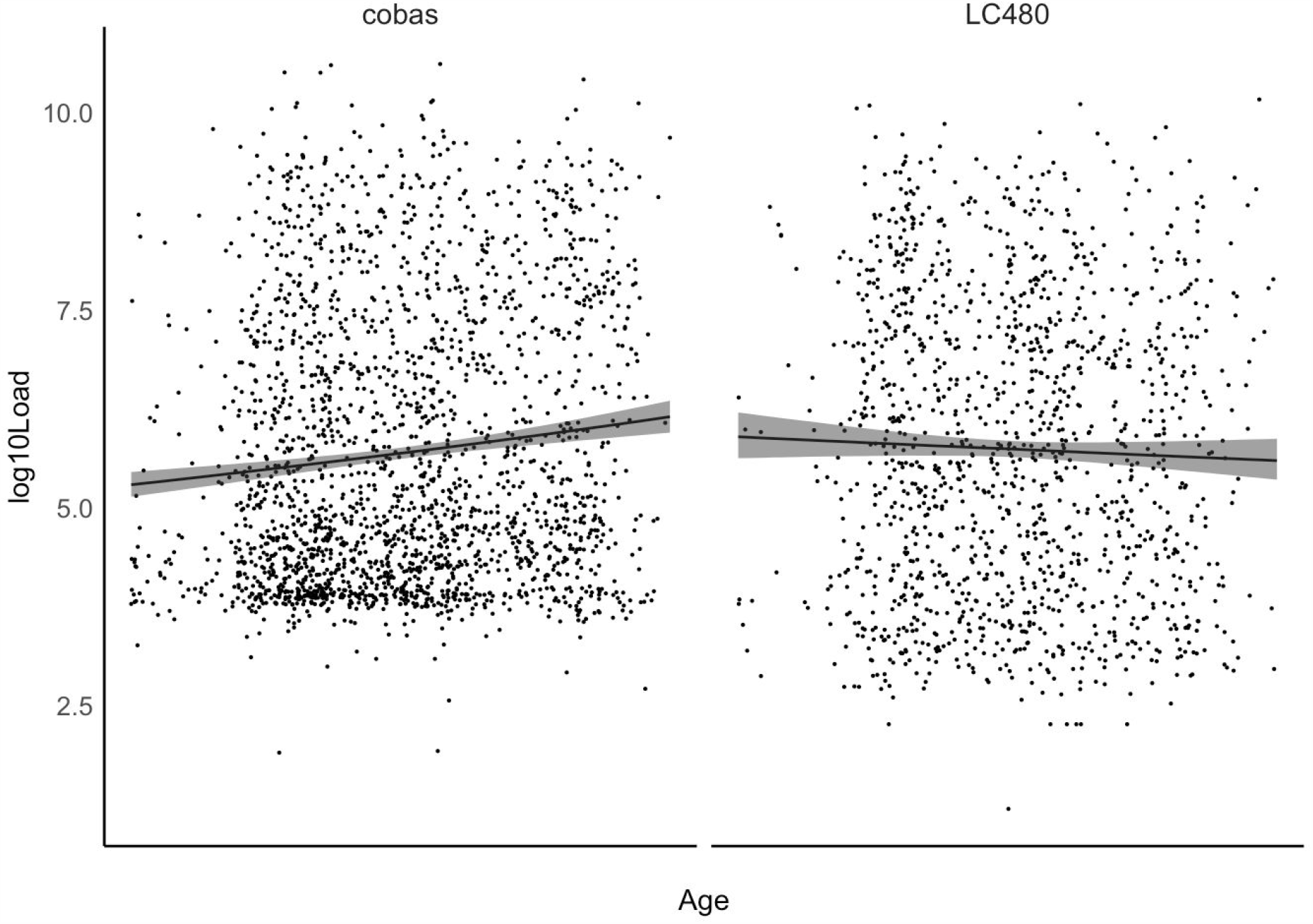
Conditional effect of age from a Bayesian gamma regression predicting viral load from age, while adjusting for type of PCR system (LC480 or cobas). Points are observed log 10 viral load, lines indicate expected log 10 viral load from the regression model. The shaded area shows a 95% credible interval. X-axis age range is approximately 0 to 100 years (exact range obscured for data privacy).

An interpretation of the difference in age/viral load regressions between LC480 and cobas is possible in light of knowledge of test utilization. For reasons stated above, children tested on cobas systems are predominantly hospitalized cases that usually have lower viral loads in throat swabs because hospitalization generally occurs after the first week of symptom onset.

Community testing on the cobas system was primarily conducted on adults as these machines were used to serve the Charité community testing and travel medicine centres which mainly see adults (**Sketch 1**).

### Percentage of patients with putatively infectious viral loads by age group

We previously established a viral load threshold of 250,000 copies per mL as the threshold for the isolation of infectious virus in cell culture at more than 5% probability (*8*). To study the percentage of people with such a viral load we used the combined data from the two PCR instruments because the data artifact in the cobas system, present in the region of viral log_10_ load 3.5 to 5.0 (**Figures 4** and **6**), does not intersect the threshold value of 5.4 (the base-10 logarithm of 250,000). Viral loads at the threshold level were present across the study period in 29.0% of kindergarten-aged patients 0-6 years (n=38), 37.3% of people aged 0-19 (n=150), and in 51.4% of those aged 20 and above (n=3153). The ∼30% figure in the young was quite similar in March (27%), April (29%), and May (30%) but due to small per-month sample numbers and the resultant inevitably broad confidence intervals, we make no statistical claims regarding these figures. Threshold percentages for a 10-year age bracket breakdown are given in **Table 2**.

## Discussion

Whereas the attack rate in children appears to correspond to that in adults (*2–5*), it is obvious that children are under-represented in clinical studies and are less frequently diagnosed due to mild or absent symptoms. For instance, a systematic review identified only 1065 pediatric SARS-CoV-2 cases in the medical literature as of April 2020 (*14*). Further, an estimate based on the number of confirmed symptomatic admissions in a specialist pediatric hospital suggested approximately 1105 (95% CI: 592-1829) unknown or unconfirmed cumulative pediatric COVID-19 hospitalizations had occurred prior to the lockdown in Wuhan starting January 23^rd^, at which point only 425 confirmed cases had been reported across all age groups, none of which were under age 15 (*15*). Because children are mostly asymptomatic, they may not be presented at testing centres even if they belong to households with a confirmed index case.

There are many other factors that complicate the determination of infection rates in children, and transmission rates from them. For instance, early transmission clusters in many countries were started by travellers of adult age, making children less likely to be index cases in households (*7*). In Germany, the fraction of children 0-19 years of age in notified new cases has almost doubled from week 12 to week 21 (March 16 to May 18), 2020, now ranging at ca. 12% of all cases (*16*). Another circumstance making children less likely to carry the virus into households is that kindergartens and schools were closed early in the outbreak in Germany, effective in most federal states from week 12 (March 16), 2020. These combined effects will cause children to be more likely to receive rather than to transmit infections in households, for purely circumstantial reasons (*7*). This phenomenon may be misinterpreted as an indication that children are less infectious.

In sum, the existing data from surveillance and observational trials show a lack of transmission data based on children. Given this, we are attempting to provide a laboratory-based proxy of infectivity based on viral load data from one of the first active testing laboratories in Germany.

Viral load is an elementary parameter in clinical virology that provides an important opportunity to directly observe viral infection. It is established that viral load correlates with infectivity on a fundamental level because the ratio between detected viral RNA and infectious virus, as measurable by virus isolation in cell culture, undergoes little variation before the onset of an adaptive immune response in acute viral diseases. The correlation between viral load and cell culture infectious dose is so well established that sensitivity limits of RT-PCR, which detect RNA, are often expressed in the form of a concentration based on tissue culture infectious doses. Our own laboratory and others have clearly shown that there is a minimum viral load, expressed in RNA copies per volume of clinical sample, beyond which clinical samples actually contain infectious virus in SARS-CoV-2 patients (*8, 17, 18*). Here, we have calculated the fractions of patients in different age strata with a sample exceeding this threshold. These figures range from ca. 30% to ca. 50% of patients in different age groups, based on data combined from LC480 and cobas testing. It is important to note that these fractions undergo a bias due to test utilization, as described, resulting in an underestimation of viral loads in the younger age groups due to representational differences that affect cobas data, and which numerically dominate the dataset when all data are analyzed together. We refrain from statistically comparing these data. Rather, we propose that ca. 30% to 50% of all swab samples that tested positive in a large laboratory contain virus concentrations that would yield virus isolates in cell culture.

Although our present knowledge of the linkage between viral load and infectivity is limited, it is incorrect that viral loads are uninformative or of unclear value, as suggested by Kaufman et al. (*19*). For instance, viral load courses in COVID-19 clearly resemble those in influenza, with both infections showing peak viral loads around the day of symptoms onset, start of virus shedding ca. 2 days prior to onset, and cessation of upper respiratory tract infectious shedding within ca. one week of onset (*8, 20–22*). For influenza, viral load-based shedding models have been established to explain the time of transmission based on dates of onset of symptoms in recipients (*20*). In influenza, the clear but imperfect correspondence between transmission timing and viral load courses indicates that viral load is relevant but unlikely to be the only factor determining transmission. This is likely also the case for SARS-CoV-2. Irrespective of this limitation, the important resemblance of shedding kinetics between SARS-CoV-2 and influenza allows comparisons that can at least provisionally inform the evolving working hypotheses in public health practice. For instance, in a study based on household contact testing for influenza (*20*), a difference in viral load of ca. 0.7 log_10_ on the day of symptom onset was associated with an increase of infectivity of 22% (i.e., 22% more secondary infections from an index case). An additional increase of 0.88 log_10_ in viral load caused infectivity to increase by another 22%. One might consider comparing these values to the difference we found between children aged 0-9 years and the older people in the cobas dataset (ca. −0.6 to −0.8 log_10_), and translating to correspondingly less infectivity based on the influenza findings. But, as explained, the LC480 dataset better represents children sampled in community/cluster testing, and these children would represent those attending kindergartens and schools. No statistically significant differences between children and adults were detected in the analyses of the LC480 dataset and the Bayesian analysis of the same dataset showed smaller differences between all groups examined than in the cobas sample. We propose that it would be incautious not to place more weight on the information from the LC480 data, and therefore assume that children of both age tiers (0-9 and 0-19 years) have virtually the same average viral loads as adults.

It is important to note that in household contact studies of influenza virus in which strict sampling and viral load testing was applied, viral loads in children and adults are not statistically significantly different (*20, 22*). Nevertheless, children with influenza H1N1 infection (141 index cases) were 2.88-fold more infectious in households relative to adults (*20*). Other differences in viral loads that are not amenable to single point measurements (as criticised by Kaufman et al. (*19*)), such as a potentially slower decline of viral load in children as compared to adults, are taken into account in these models, pointing to other explanations for higher infectivity in children, such as contact frequency and intensity (*20*). It was shown that age-specific behavioural differences make a large contribution to the established higher infectivity of children compared to adults in influenza, which is an important consideration given the lack of knowledge on SARS-CoV-2 transmissibility (*23*). As pre- or mild-symptomatic behavioural traits are likely virus-independent, the viral load results from the present study cannot be ignored in discussions on potential infectivity. An unlimited re-opening of kindergartens and schools would re-establish behavioural traits that facilitate virus transmission through contact. Based on the example of influenza, where similar viral loads in children and adults coincide with an increased role of schools and kindergartens for the maintenance of epidemic waves, the unlimited opening of these facilities should be carefully monitored by preemptive diagnostic testing.

The meaning of symptoms for actual transmissibility of SARS-CoV-2 is unclear. For influenza there is evidence suggesting that severity of symptoms predicts transmissibility. In one study on household clusters, index cases with fever were 94% more infectious than those without (*20*). For influenza, symptoms in turn seem to correlate with viral load. For instance, the difference in viral load between symptomatic and paucisymptomatic cases in another study was 1-2 log_10_, increasing with time from onset of symptoms, after being very similar at onset (*24*). However, this correlation may not exist in SARS-CoV-2 infection, which means that the absence of symptoms does not necessarily imply lower levels of virus excretion. In a study of people living in the Italian village of Vó, in which ca. 80% of the population were tested by RT-PCR twice within two weeks, about half of the population were found to be asymptomatically infected, showing no symptoms over the entire observation period of two weeks, while viral loads were equivalent in symptomatic and asymptomatic patients (*25*). This observation corresponds to our data in children, in particular for the LC480 dataset that contains more children who are likely to present no or mild symptoms.

It is a limitation, due to the urgency and workload of the pandemic context, that metadata needed to discriminate patients into sub-cohorts based on symptomatic status, underlying diseases, or other indications for diagnostic test application, is not available. However, for 47 cases (1-11 years of age) for whom information on diagnostic indications was available, we identified fifteen cases with indications of underlying disease or hospitalization. These children had lower viral loads than those without known underlying disease tested in outpatient departments, practices, or households (**Figure 2**). The latter set represents children more likely to be attending schools and kindergartens.

### Influence of symptoms on SARS-CoV-2 detection

In the LC480 data that better represent community/cluster testing, the virus detection rate increased steadily within the younger age groups of patients tested, before reaching a plateau from middle-aged adults and older (**Table 2**). As testing was predominantly directed by symptoms, this may be explained by the fact that the value of symptoms to inform diagnostic testing increases over the first half of life. The clinical specificity of laboratory diagnostics in community testing may be lower for children than for adults (**Sketch 2**). This is because children with respiratory symptoms and fever will be less likely to have an acute SARS-CoV-2 infection than adults with similar symptoms. Many other respiratory viruses cause symptomatic disease with fever in children and young adults, but less so in adults where endemic respiratory viruses often present as mild upper respiratory tract infections without fever, a condition that often triggered laboratory testing in adults, particularly in the first half of the observation period. Our results should clearly not be taken as an indicator of age-specific prevalence in Germany. Rather, the low rate of SARS-CoV-2 detection confirms that symptoms are not a good predictor of infection in children. This is an additional challenge in monitoring kindergartens and schools upon unlimited re-opening. Intense sentinel testing by RT-PCR may be necessary to ensure early outbreak detection in absence of symptoms.

The viral loads observed in the present study, combined with earlier findings of similar attack rate between children and adults (*2–5*), suggest that transmission potential in schools and kindergartens should be evaluated using the same assumptions of infectivity as for adults. There are reasons to argue against the notion of adult-like infectivity in children, such as the fact that asymptomatic children are less likely to spread the virus by coughing, and have smaller exhaled air volume than adults. However, there are other arguments that speak in favour of increased transmission probability, such as the greater physical activity and closer social engagement of children. We recommend collecting and evaluating more viral load data from testing laboratories to achieve more robust statistical assessments and independent confirmation of the present results.

## Data Availability

The data will be made available upon publication.

## Acknowledgements

Work at Charité virology is funded by European Commission via project ReCoVer, the German Ministry of Research and Education via Deutsches Zentrum für Infektionsforschung, and the German Ministry of Health via the Konsiliarlabor für Coronaviren.

## Methods

### Data curation and anonymization

Research clearance for the use of routine data from anonymized subjects is provided under paragraph 25 of the Berlin *Landeskrankenhausgesetz*.

All data are anonymized before processing to ensure that it is not possible to infer patient identity from any processing result. The anonymization procedure is as follows. Patient names are replaced with a value from a secure one-way hash function. Patient ages (in days) are modified by the addition or subtraction of a random value drawn from a normal distribution, and rounded before assignment to age categories. Sample collection and processing dates are similarly randomly adjusted. RT-PCR threshold cycle values (received by us with only one decimal place of precision) are used to estimate viral load (limited-precision conversion formulae are given below) and these values are then also randomly adjusted. Age and timeline details are omitted from the x-axis in figures in this manuscript and dates of sample collection are not indicated. Patient health status for patients whose data are shown in Figure 2 is divided into two categories: a) a single pooled group, with no indication of actual status, of those who are hospitalized or with a pre-existing condition, and b) all others. Samples collected from test centres preferentially seeing those with early and mildly symptomatic cases (community/cluster testing) are pooled to create a single binary category, again with no information retained regarding test centre identity. A similar anonymized binary pooling is used to distinguish between samples collected at Charité test centres and samples collected elsewhere. All randomization was performed using the Mersenne Twister algorithm (with period 2^19937^-1), as implemented in the `random` module of Python (version 3.8.2). The seed used to initialize the random number generator was not pre-specified and the automatic, internally-generated, key was not output or otherwise retained.

Due to testing of some but not all positive cases by two RT-PCR targets, 3303 of 77,996 (4.23%) patients had 7,032 positive results overall. In cases with more than one result, we selected the first (i.e., earliest) RT-PCR result. Almost all testing relied on naso- and oropharyngeal swab samples. Less than 3% of all samples were samples from the lower respiratory tract, and these were not removed from the dataset because of their low number and the fact that first samples per patient almost universally are swab samples (samples from the lower respiratory tract are generally taken from patients only after intubation, by which point viral loads have typically fallen).

### Viral load analysis

The viral load projection derived in our study is semi-quantitative, and projects viral load per mL of sputum or per entire swab sample, while only a fraction of the volume of both types of sample can actually reach the test tube. Also, quantification is based on a standard preparation tested once in multiple diluted replicates to generate a standard curve and derive a formula upon which Ct values are transformed into viral loads. This approach does not reflect inter-run variability or the variability between different RT-PCR setups and chemistries. However, these variabilities apply to all age groups and do not affect the interpretation of data for the purpose of the present study.

The following Python (version 3.8) software packages were used in the analysis and production of images: Scipy (version 1.4.1) (*26*), pandas (version 1.0.3) (*27*), statsmodels (version 0.11.1) (*28*), matplotlib (version 3.2.1) (*29*), numpy (1.18.3) (*30*), seaborn (version 0.10.1) (*31*), and scikit_posthocs (version 0.6.4) (*32*).

Viral load is estimated from Ct value based on the empirical formulae log_10_(1.441E14 * exp(−0.685 * ct)) for the LC480 system and log_10_(1.105 * exp(−0.681 * ct)) for the cobas system. The formulae are derived from testing standard curves. The exact constant values used are here truncated to three decimal places.

The Bayesian analysis of viral load data used gamma likelihood functions to account for the restriction of outcome values to positive numbers. The primary analysis used a mixture of three gamma distributions in order to account for the multi-modality of the log_10_ viral load values. The analysis that used age as a continuous outcome variable used a standard gamma likelihood function which does not capture multi-modal outcomes in this analysis. All Bayesian models used weakly informative priors and were estimated using 4 chains with 1000 warm-up samples and 1000 post-warm-up samples. Convergence of MCMC chains was examined by checking that all Potential Scale Reduction Factors (R-hat) values were below 1.1.

## Informal sketches

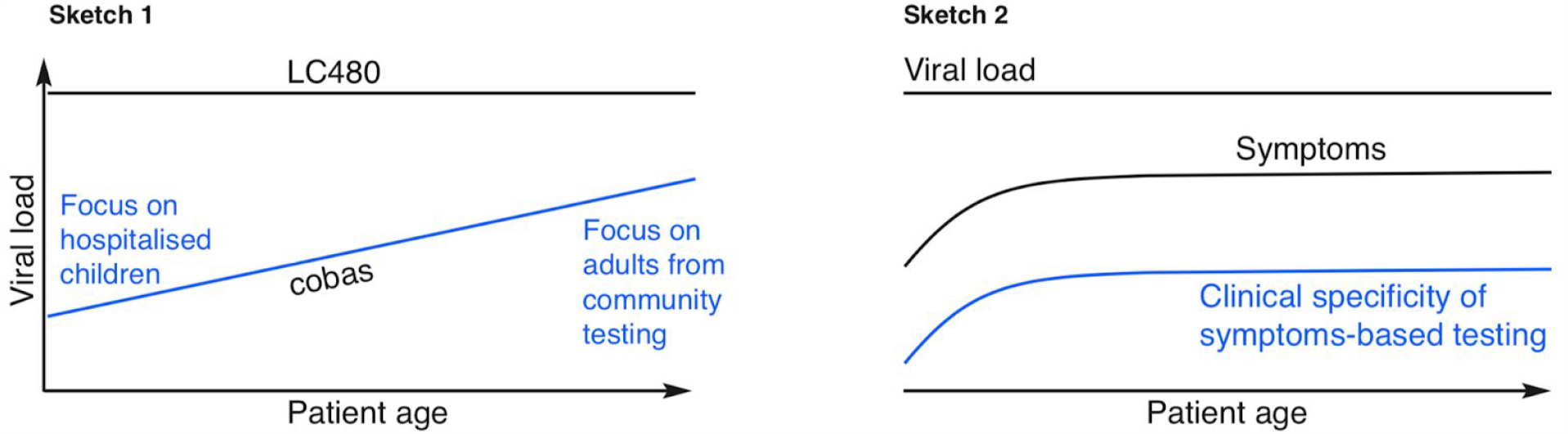

### Sketch 1

Qualitatively illustrates the change in viral load for the cobas system according to patient age. Due to the required use of a specific sample buffer, cobas systems were typically only used in contexts involving close collaboration between Labor Berlin and medical centres (e.g., the Charité community testing centre and the Charité travel medicine outpatient department) where patient composition is biased towards older people. Children who are hospitalised and those with underlying medical conditions are disproportionately more likely (compared to other children) to be tested on cobas systems and the time delay involved in their testing (typically ca. one week) results in lower viral loads (**Figures 2** and **3**).

### Sketch 2

Qualitatively illustrates how the clinical specificity of symptoms-based testing increases with patient age. This is because children with respiratory symptoms and fever will be more likely to have a non-SARS-CoV-2 infection than adults with similar symptoms. Many other respiratory viruses cause symptomatic disease in children and young adults, but less so in adults where endemic respiratory viruses often present as mild upper respiratory tract infections without fever. Thus younger people sent for SARS-2-CoV diagnostic testing on the basis of symptoms are less likely to actually have that infection. In older people, the range of likely causative pathogens is reduced, resulting in a higher SARS-2-CoV detection rate in symptoms-directed testing.

### Reply to collective criticism of statistical methodology in the initial preprint

Consistent and constructive feedback was received regarding the statistical analysis of the original preprint. It was uniformly pointed out that dividing the data into ten categories and doing an all-pairs comparison invokes many (socially and practically) irrelevant comparisons, and that this division also reduces power by creating sub-groups with small cardinality. It was also pointed out that the Kruskal-Wallis test with post hoc analysis could be omitted. For completeness we had included the results of the Tukey HSD and Bonferroni-adjusted t-tests but should not have, and our description of the results should not have referred to them. It was also suggested that we additionally treat age as a continuous variable in an overall regression analysis. To address these issues, we now examine just three divisions of the samples (0-9 years versus >9 years, 0-9 versus >19, and 0-19 versus >19). These divisions are more socially relevant and, other than the 0-9 years group, have higher cardinality. We followed the suggestions to drop the Kruskal-Wallis test and 45 post hoc pairwise tests and instead use Welch’s t-test and the Mann-Whitney rank test to examine the three binary splits just mentioned. We added a Bayesian analysis of the differences between these categorizations and a Bayesian analysis with age as a continuous variable (based on day of birth) predicting log_10_ viral load. Importantly, we detected an unexplained artifact in the Ct values reported by the cobas instrument. Future work is planned to quantify and possibly correct for the impact of this artifact. For now, we have separated the analysis of the two instruments (except where considering presumably unaffected higher viral loads and the 250,000 viral copy threshold). We have also investigated the differential use of the two instruments due to various social and professional behavioural factors and above discuss how these issues are important for the interpretation of the raw data. We now also provide discussion on the relationship between formal statistical significance (including quantitative estimates of differences between estimated viral loads) and possible clinical significance.

